# External Validation and Comparison of Two Clinical Prediction Models (PTP2013 and PTP2019) for Chest Pain in a Colombian Cohort

**DOI:** 10.1101/2025.11.05.25339632

**Authors:** Nathaly Puentes, Carlos Daniel Rodríguez-Ariza, Federico Ramos-Marquez, Diego Alejandro Vargas-Hernández, Sergio Moreno, Laura Sofia Guevara, Laura Daniela Rincón, Karen Morales, Santiago Vargas Paredes, Santiago Callegari, Carlos Andrés Sánchez-Vallejo

**Author notes:** Corresponding author: Federico Ramos.

## Abstract

**Aims:** The European Society of Cardiology (ESC) has proposed four pre-test probability (PTP) models for obstructive coronary artery disease (CAD). However, no studies have evaluated the diagnostic performance of any predictive model in the Latin American population. The aim of this study is to compare the PTP2013 and PTP2019 predictive models in order to determine which demonstrates a superior diagnostic performance for CAD in a cohort of Colombian patients.

**Methods:** A total of 408 patients who presented with chest pain and underwent coronary angiography (CA) and/or coronary computed tomography angiography (CCTA) at Fundación Santa Fe de Bogotá, between January 2019 and December 2023 were enrolled. Medical records were retrieved from the Hemodynamics and Radiology units. Pre-test probabilities were calculated for each patient using both the PTP2013 and PTP2019 models. CAD was defined as >50% stenosis on CA or CCTA. Each predictive model was assessed against CA and/or CCTA findings. The comparative performance of both models was evaluated.

**Results:** Prevalence of obstructive CAD of 24.9%. The PR2019 model underestimated the probability of CAD by 59%, whereas the PTP2013 model overestimated it by 35.6%. PTP2019 model yielded a C-statistic of 0.610 [95% CI: 0.544 - 0.676], while the PTP2013 model reported a C-statistic of 0.633 [95% CI: 0.570 - 0.696] (comparative p-value: 0.060). The net reclassification improvement was 14.7%). At a 15% threshold, the PTP2013 model demonstrated a sensitivity of 90% (82.38 - 95.10%), compared to 48% (37.9 - 58.22%) for the PTP2019 model.

**Conclusion:** The PTP2013 model is favored, as it showed higher sensitivity and a tendency to overestimate risk, in contrast to the PTP2019 model, which exhibited a concerning underdiagnosis of CAD. Consequently, the methodological challenge of identifying the predictive model with the highest diagnostic performance remains, highlighting the need to develop a tailored prediction model for the local population.

## Introduction

Cardiovascular disease is recognized as the leading cause of morbidity and mortality within non-communicable diseases worldwide (Roth et al., 2018). In 2019, 197 million prevalent cases of coronary artery disease (CAD) were reported worldwide (Roth, Mensah, Johnson, et al., 2020), and 9.4 million deaths were reported in 2021 (Vaduganathan et al., 2022). Therefore, early and timely detection of CAD is a fundamental pillar; consequently, predictive models for CAD have been developed.

To effectively diagnose CAD, the calculation of a clinical prediction model is essential. Predictive models, known as pretest probability testing (PTP), have experienced a gradual decline in the estimation of the probability of CAD. This significant shift is attributed to a substantial reduction in the prevalence of CAD (Juarez Orozco et al., 2019). The initial revision of the original Diamond Forrester PTP was conducted in 2013, based on a 49% prevalence of CAD. Conversely, the 2019 PTP was updated based on a prevalence of only 14.9%. It is noteworthy that these data originate from European and North American cohorts (Juarez-Orozco et al., 2019 & Winther et al., 2022), excluding the Latin American population.

On the other hand, the prevalence of CAD in Colombia remains undisclosed. However, it is documented that 80% of global cardiovascular mortality originates from developing and low-income countries (Roth et al., 2020). Furthermore, data on cardiovascular risk factors in Latin America are extensively reported (Roth, Mensah, & Fuster, 2020). Consequently, the clinical and epidemiological characteristics of the Latin American population exhibit distinct differences from those of the European population. Consequently, it is imperative to validate the models at the local level.

Based on the above, the decrease in the prevalence of CAD led the European cardiology guidelines to adjust the most recent prediction model. Among the multiple existing prediction models, the models recommended by the European cardiology guidelines are: the PTP 2013 (Montalescot et al., 2013) and the PTP 2019 (Neumann F. J., 2019), algorithms or models that have the most evidence available and their validation process has been developed in different populations (Genders et al., 2011, 2012 & Juarez et al., 2019). Although both models consider the same clinical variables (age, sex, and type of chest pain), and the same probability categorization (low, intermediate, high), with respect to PTP2013, the PTP2019 model presented a change in the probability calculation.

The categorization of PTP2013 and PTP2019 holds significant importance in determining appropriate medical interventions based on the likelihood of CAD occurrence. Patients classified as intermediate or high probability undergo functional or invasive cardiac testing, while those classified as low probability receive expectant management without medical intervention. (Neumann F.J et al., 2019). The PTP2019 update resulted in a 50% reclassification of patients from intermediate to low probability compared to PTP2013. (Juarez et al., 2019) However, the absence of diagnostic validation of these prediction models in the Colombian population introduces uncertainty. Classifying a larger proportion of patients in the low probability group may increase the risk of false negatives and subsequent under-diagnosis of CAD.

Recent external validation studies conducted in China have substantiated the underestimation of the probability of PTP 2019 compared to PTP 2013 (Zheng et al., 2024). In light of these findings, it is imperative to conduct the external validation of the clinical prediction models PTP2013 and PTP2019 in the Colombian population.

The objective of the present study is to assess and compare the diagnostic performance of both models (PTP 2013 and PTP2019) within a Colombian cohort of patients presenting with chest pain. The outcomes of this study will facilitate the identification of the most effective model for predicting coronary artery disease (CAD) in the Colombian context. These results will enable medical personnel in primary care centers to recommend the most accurate model, thereby guiding clinical decisions and facilitating a practical and timely approach to the identification of CAD. The optimal diagnosis will facilitate early intervention for referral to medical or invasive management, thereby minimizing the risk of medium and long-term complications associated with CAD. Furthermore, these findings will provide evidence of the predictive performance of PTP2013 and PTP2019 in a local setting, paving the way for future research in this area.

## METHODS AND MATERIALS

This is a retrospective observational study based on diagnostic tests. Data were collected from the Hemodynamics and Radiology databases of patients who underwent coronary arteriography or coronary CT angiography between January 2019 and December 2023 at the Hospital Universitario Fundación Santa Fe de Bogotá (FSFB).

### Participants

Colombian patients over 18 years of age who presented with chest pain and/or dyspnea and underwent coronary CT angiography and/or coronary angiography were included. Patients were excluded if they had suspected acute coronary syndrome, were undergoing pre-surgical evaluation, had heart failure with an ejection fraction <50%, dyspnea classified as functional class III or IV, structural heart disease, or a history of coronary artery disease or coronary revascularization. The sample size was determined using consecutive non-probability sampling and was calculated for a diagnostic test study, assuming a prevalence of heart disease of 53%, a 95% confidence interval, a 20% margin of error, and a cutoff point of 15%, yielding a minimum required sample size of 391 patients. Statistical analyses were performed using Stata 18 software.

### Index or Probability test

The index test was evaluated with two clinical prediction models, each model calculates the likelihood of a patient presenting with CAD, the probability calculation is based on clinical characteristics, without including invasive procedures. The prediction models were based on logistical regression models to predict the likelihood of CAD. These algorithms evaluate the likelihood of CAD occurence: these are practical predictive models as they do not require invasive procedures. Both have been previously validated in European and North-American cohorts. The selected models are the Pre Test Probability model of Diamond-Forrester updated in 2013 (annex 1) and Pre Test Probability model of 2019 (annex 2). Both take into consideration the following variables: age, sex, and characteristics of chest pain; the ESC-PTP 2019 model includes dyspnea. Chest pain was defined in three categories: typical angina, which meets three criteria: oppressive retrosternal, caused by physical or psychological stress, and alleviated by rest and/or nitrates; atypical angina which meets two out of three criteria listed above; and non-coronary chest pain, which meets one or none of the criteria above (Montalescot et al., 2013b).

Both the ESC-PTP 2019 and ESC-PTP 2013 models state the probability in a scale from 0 to 100 and are categorized according to european guidelines (Neumann et al., 2020).

For bias assessment, patients with missing data for index test (PTP 2013 and PTP 2019) were excluded; besides, researchers who registered the index test had no knowledge of the reference standard.

### Standard reference

The standard reference was coronary arteriography (CA), which was defined as an obstruction greater than 50% of a main coronary vessel. To accomplish an evaluation that includes the totality of the probability spectrum of a coronary obstruction, the coronary angiotomography (Angio-CT) is also considered an appropriate alternative to standard reference, given it has an excellent sensibility (96-100 %) and specificity (85-92 %) according to previous studies (Cury et al., 2016).

CA was performed through radial or femoral access, the procedure was completed according to standard medical practice measure, with a minimum of 12 views. Images were evaluated by two interventional cardiologists. The inter-observer differences were solved by consensus between them and had no access to the prediction models. CA was not performed in all patients, considering this procedure was conducted in patients previously considered with intermediate and high probability poor CAD as per PTP 2019 (Fig. 1). The information was registered in medical records after the procedure was done by the cardiologist.

Angio-CT were performed using a 64-slice dual-source CT scanner (Siemens Medical Systems). The severity of the disease was defined based on the greatest stenosis identified among all evaluated segments. Radiologists did not have access to model prediction results either.

### Ethical considerations

In accordance to Resolution 8430 of 1993 issued by the Colombian Ministry of Health, this study is classified under the minimum risk category, as it does not involve medical interventions or invasive procedures, and data is collected from registries of medical records as established in the Declaration of Helsinki – 59th General Assembly, Seoul, South Korea, October 2008. The study had investigation ethics committee aproval in FSFB under record number **APY-CCEI-F-028.**

## STATISTICAL ANALYSIS

Participants were selected based on the CA and CCTA records. Subsequently, the PTP 2013 and PTP 2019 clinical prediction models were calculated for each individual, followed by appropriate categorization for each model. Statistical analysis was conducted in three phases. First, a descriptive analysis of the total study population was performed. Then, two groups were established based on the presence of coronary artery occlusion and compared using chi-square, Student’s t-test, or Mann–Whitney U test, as appropriate. The final phase involved external validation by calculating the scores for each model (PTP 2013 and PTP 2019) and comparing their diagnostic accuracy in terms of calibration, discrimination, reclassification indices, and clinical significance.

### Calibration

Calibration is use to assess the reliability of the predicted probabilities by evaluating the consistency between predicted and observed outcomes. Initially, large-scale calibration was assessed by comparing the mean predicted proportion of obstructive CAD with the observed proportion in the sample.

The goodness of fit was assessed using the Hosmer–Lemeshow test for both PTP 2013 and PTP 2019 (Steyerberg, 2019).

Calibration was visualized using two graphs. The first was a graph of predicted against observed endpoints across a continuous probability scale from 0 to 100. In a perfectly calibrated model, the calibration curve has a slope of 1 and an intercept of 0, forming a 45-degree line. The observed calibration curve was interpreted as follows: if the line lies below the diagonal, it has a negative value and indicates overestimation; if it lies above, it indicates a positive value and underestimation of the model (Steyerberg & Vergouwe, 2014). A comparative calibration plot of both models was constructed.

The second graph consisted of bar charts stratified by probability quartiles, visually comparing observed versus expected prevalence for each model.

### Discrimination

Discrimination refers to the model’s ability to distinguish individuals with obstructive CAD from those without it. It was assessed using the C-statistic (Cook, 2007). For binary outcomes, the C-statistic is equivalent to the area under the receiver operating characteristic curve (AUC or ROC). Sensitivity was plotted on the Y-axis and 1-specificity on the X-axis, resulting in a curve that illustrates how sensitivity varies with specificity as the decision threshold shifts across a 0–100 probability scale.

ROC curves were plotted for both models on the same graph. Higher values (closer to 1) indicate better predictive performance. Effect size was interpreted following Collen’s methodology (Rice et al., 2005).

Finally, areas under the ROC curves were compared using the non-parametric DeLong method to determine which model had superior discriminative power, reported with the corresponding confidence intervals (Pencina et al., 2008). The DeLong test evaluates the null hypothesis that both models have equal discriminative ability; thus, a p-value < 0.05 suggests a statistically significant difference between the ROC curves.

### Net Reclassification Improvement (NRI)

NRI was used to quantify how many patients were correctly reclassified by the updated PTP 2019 model, both among those with confirmed CAD and those without disease. Each patient was initially classified into one of three risk categories: low, intermediate, or high. The number of patients with CAD who were correctly reclassified to higher-risk categories, as well as the number of patients without CAD who were correctly reclassified to lower-risk categories, was determined. The total NRI was calculated as the sum of both improvements. A value of 0 indicates no difference between models; a value >0 indicates improved classification by the new model, and a value <0 favors the older model.

### Clinical Significance

Clinical significance for decision-making was assessed according to Steyerberg, E. W., by conducting a sensitivity analysis for each model. For both clinical prediction models, the low-probability category was defined as <15%. An analysis of sensitivity, specificity, positive predictive value (PPV), negative predictive value (NPV), positive likelihood ratio (LR+), negative likelihood ratio (LR−), and diagnostic odds ratio (DOR) was obtained.

### Verification Bias Correction

Since the reference standard (CA) was applied only to a subset of patients (those initially categorized as intermediate or high risk by the PTP 2019 model), it was necessary to include the full disease spectrum to avoid verification bias. First, a propensity score was calculated for each selected patient using a logistic regression model to estimate the probability of undergoing CA, based on predictive variables such as age, sex, and chest pain characteristics. Inverse probability weighting (IPW) was then applied by assigning lower weights to higher propensity scores. This process was applied to each individual. Lastly, a logistic regression model was used to estimate the probability of obstructive CAD based on sex, age, chest pain type, and the IPW-adjusted weights.

## RESULTS

### Study Population

During the study period, a total of 1,677 patients were referred for coronary angiography (CA) and coronary computed tomography angiography (CCTA). Following the application of exclusion criteria, 361 patients were included in the final analysis. Among the excluded subjects, the absence of a complete medical record was the primary reason for exclusion, accounting for 490 patients. A total of 408 patients were selected, of whom 285 had undergone CA and 85 CCTA. Among the selected patients who had CA, 70% had significant coronary artery disease (CAD), defined as >50% obstruction of a main vessel. In the CCTA group, 64% were found to have significant CAD.

The validation cohort included 408 patients (51.7% women), of whom 300 had CA and 135 underwent CCTA. The proportion of subjects with documented coronary artery obstruction by arteriography and/or angiotomography was 24.51% (n = 100), with a higher frequency in men (56%) and in the 60–69 age group (33.8%).

Regarding comorbidities in the coronary obstruction group, dyslipidemia was the most prevalent risk factor, documented in 73% of subjects, with no statistically significant difference between sexes (p = 0.39). Arterial hypertension was the second most frequent comorbidity, present in 38.4% of patients, most of whom were classified as Stage I according to the Eighth Joint National Committee. Diabetes mellitus was observed in 19.8% of patients, with significantly higher glycemia levels compared to the no-obstruction group (p < 0.001). Although a history of tobacco use was more common among those with coronary artery obstruction, this difference was not statistically significant (p = 0.19). No statistical differences were observed regarding family history of CAD. Notably, 42.4% of patients had a body mass index (BMI) between 25 and 29.9, classifying them as overweight.

A total of 65.6% of patients had some degree of decreased glomerular filtration rate. Patients with an estimated glomerular filtration rate (eGFR) below 45 mL/min/1.73 m², as measured by the MDRD formula, had a significantly higher rate of coronary obstruction (p = 0.007) compared to the no-obstruction group. The majority of patients with stable angina underwent some form of non-invasive cardiac testing (n = 320); of these, coronary angiotomography was the most frequently performed (33.9%), while cardiac magnetic resonance imaging was the least used. Table 1 presents the complete demographic characteristics of the cohort.

Among patients with coronary artery obstruction, non-anginal chest pain and dyspnea were the most common clinical presentations, reported in 44% and 42% of cases, respectively. Referred pain was present in only 27% of patients. Furthermore, among individuals with coronary artery obstruction, only one (1) patient was classified as low probability according to the 2013 ESC-PTP model, while 89% were classified as intermediate probability. In contrast, under the 2019 ESC-PTP model, 36% of patients were classified as low probability and 62% as intermediate probability. According to the 2013 ESC-PTP model, the mean predicted probability was 44% (IQR: 24.0–50.0); in comparison, the 2019 ESC-PTP model estimated a lower mean probability of 16% (IQR: 10.0–26.0). Table 2 presents the complete clinical characteristics related to chest pain.

**Figure 1.**
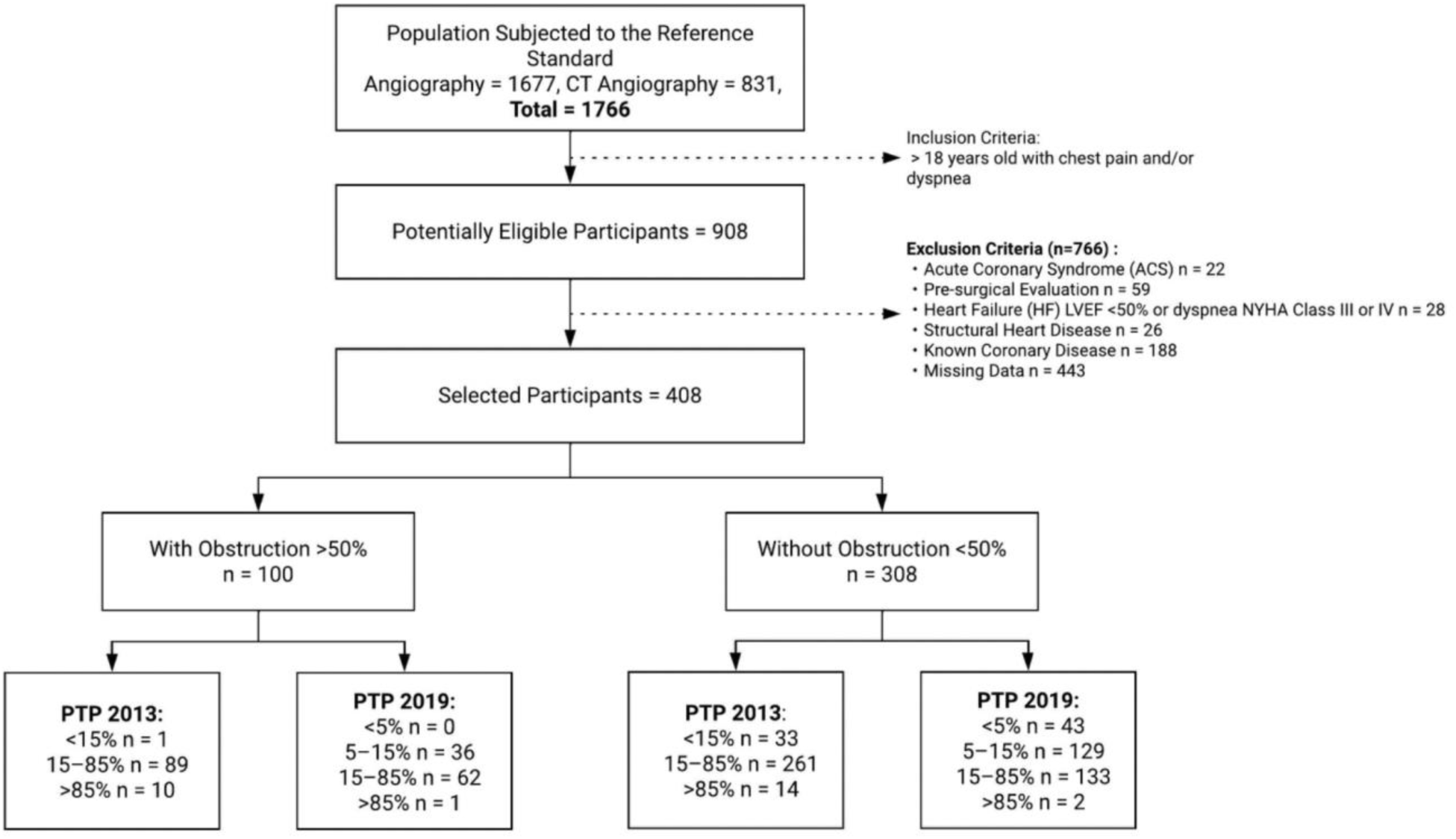
Study flow chart. PTP=pre-test probability

### Large-scale Calibration

The large-scale calibration analysis revealed significant discrepancies between the observed mean outcomes and the predicted mean probabilities, indicating poor calibration in both models. For the PTP2013 model, the calibration intercept was -0.804, with a calibration slope of 0.391. In the case of the PTP2019 model, the calibration intercept was 0.798, and the slope was 0.259.

The PTP2013 model showed a 35.6% overestimation overall, which was particularly pronounced in women aged 40 to 49, where the predicted probability was 41.46% compared to an observed proportion of 8.7%. Overestimation was evident across all probability categories, especially in the high-probability group, where the predicted probability reached 69.9% compared to an observed proportion of 40% (Annex 1).

**Table 1.**
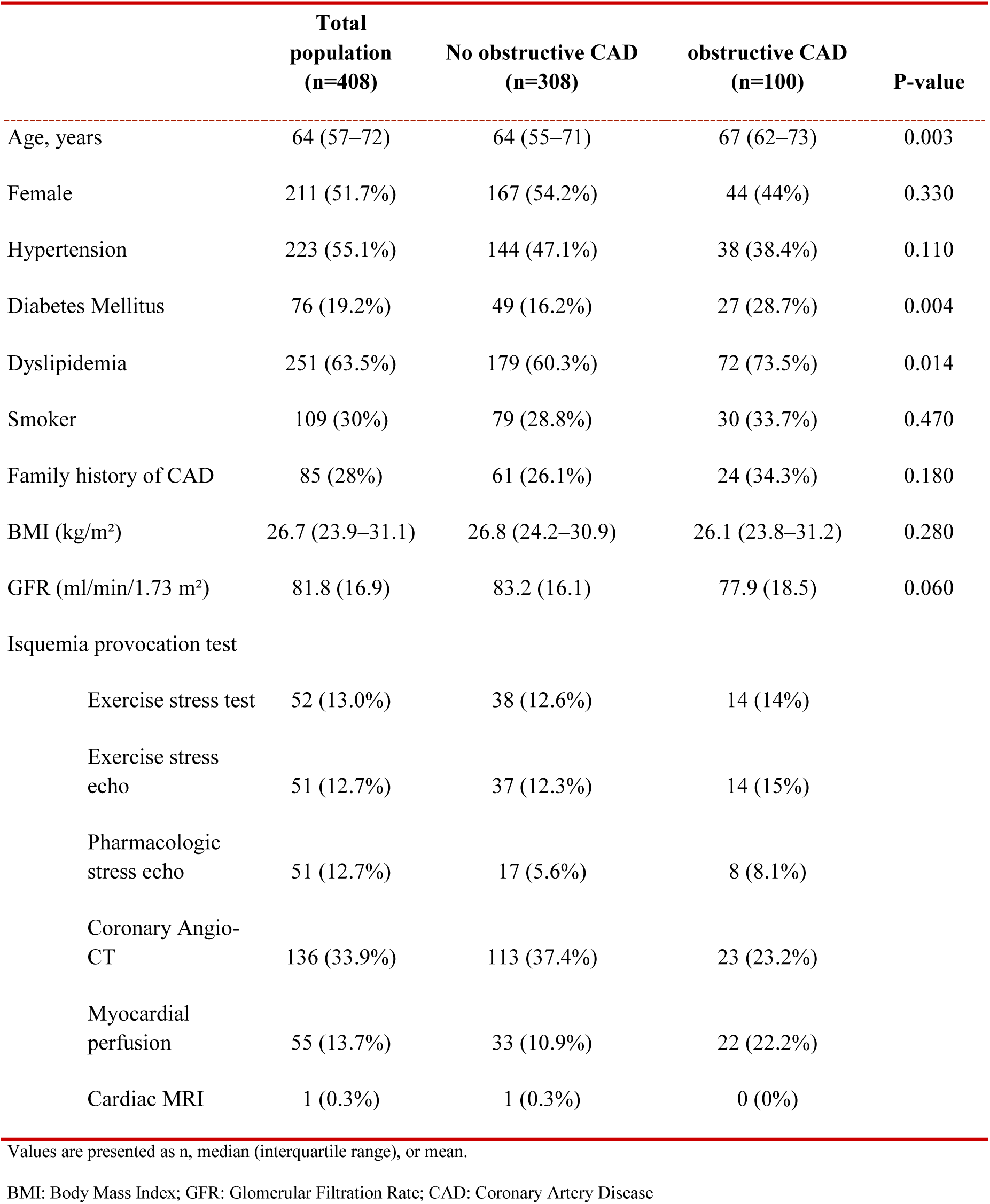
Baseline Characteristics of the Patients.

|  | **Total**<br>**population**<br>**(n=408)** | **No obstructive CAD**<br>**(n=308)** | **obstructive CAD**<br>**(n=100)** | **P-value** |
| --- | --- | --- | --- | --- |
| Age, years | 64 (57–72) | 64 (55–71) | 67 (62–73) | 0.003 |
| Female | 211 (51.7%) | 167 (54.2%) | 44 (44%) | 0.330 |
| Hypertension | 223 (55.1%) | 144 (47.1%) | 38 (38.4%) | 0.110 |
| Diabetes Mellitus | 76 (19.2%) | 49 (16.2%) | 27 (28.7%) | 0.004 |
| Dyslipidemia | 251 (63.5%) | 179 (60.3%) | 72 (73.5%) | 0.014 |
| Smoker | 109 (30%) | 79 (28.8%) | 30 (33.7%) | 0.470 |
| Family history of CAD | 85 (28%) | 61 (26.1%) | 24 (34.3%) | 0.180 |
| BMI (kg/m<sup>2</sup>) | 26.7 (23.9–31.1) | 26.8 (24.2–30.9) | 26.1 (23.8–31.2) | 0.280 |
| GFR (ml/min/1.73 m<sup>2</sup>) | 81.8 (16.9) | 83.2 (16.1) | 77.9 (18.5) | 0.060 |
| Ischemia provocation test |  |  |  |  |
| Exercise stress test | 52 (13.0%) | 38 (12.6%) | 14 (14%) |  |
| Exercise stress echo | 51 (12.7%) | 37 (12.3%) | 14 (15%) |  |
| Pharmacologic stress echo | 51 (12.7%) | 17 (5.6%) | 8 (8.1%) |  |
| Coronary Angio-CT | 136 (33.9%) | 113 (37.4%) | 23 (23.2%) |  |
| Myocardial perfusion | 55 (13.7%) | 33 (10.9%) | 22 (22.2%) |  |
| Cardiac MRI | 1 (0.3%) | 1 (0.3%) | 0 (0%) |  |

Values are presented as n, median (interquartile range), or mean.
BMI: Body Mass Index; GFR: Glomerular Filtration Rate; CAD: Coronary Artery Disease

In contrast, the newer PTP2019 model demonstrated a 59% underestimation, most notably in men aged 60 to 69, where the predicted probability was 7.57% versus an observed proportion of 28%. Underestimation was also greater in the low-probability group, with an observed proportion of 21% compared to a predicted probability of 1.57% (Annex 1).

Both models produced extreme predictions, with calibration slopes of 0.391 (PTP2013) and 0.259 (PTP2019), values that deviate substantially from the ideal slope of 1 (Figure 2).

**Figure 2.**
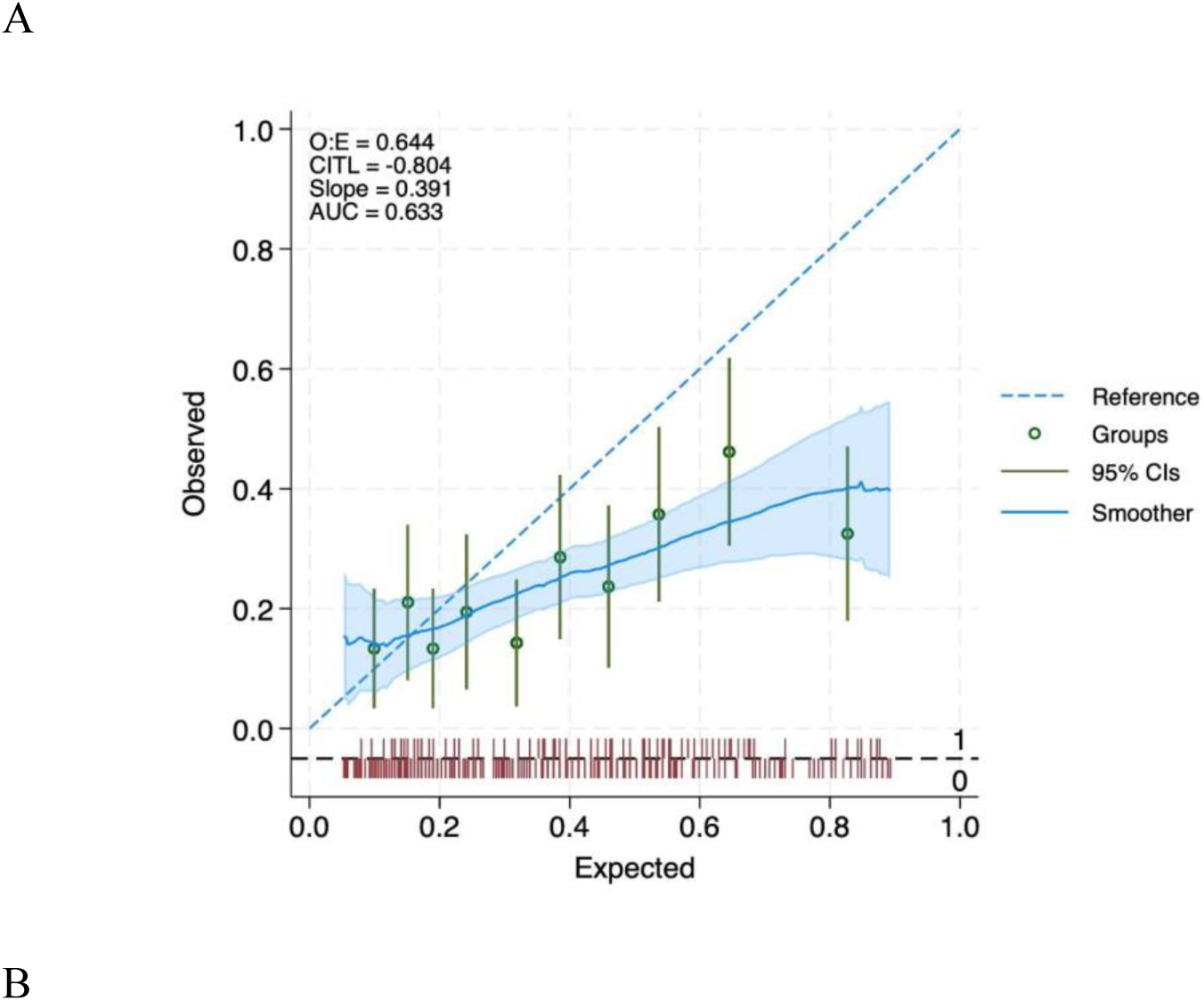
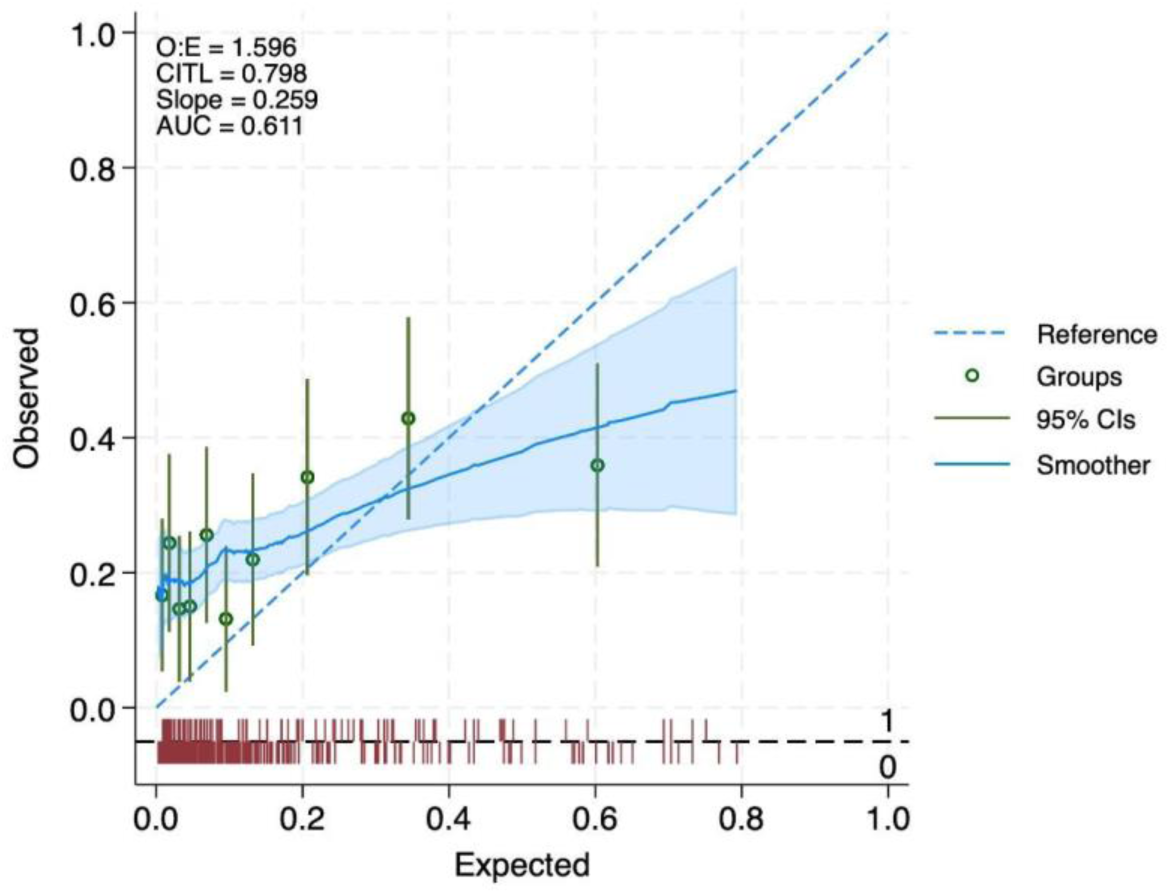
Calibration Plots for the 2013 and 2019 PTP Models in Predicting Coronary Artery Disease.The intercept and slope are shown. The ideal 45-degree line has an intercept of 0 and a slope of 1. Circles represent the results for deciles of predicted risk, with 95% confidence intervals for the observed proportions of Coronary Artery Disease.

**Table 2.** Patient pain characteristics.

|  | Total<br>population<br>(n=408) | No obstructive CAD<br>(n=308) | Obstructive CAD<br>(n=100) | P-value |
| --- | --- | --- | --- | --- |
| **Symptoms** |  |  |  |  |
| Typical chest pain | 83 (20.3%) | 56 (18.2%) | 27 (27%) | 0.16 |
| Atypical chest pain | 133 (32.6%) | 104 (33.8%) | 29 (29%) | 0.16 |
| Non-anginal pain | 192 (47.1%) | 148 (48.1%) | 44 (44%) | 0.16 |
| Dyspnea | 162 (39.7%) | 120 (39%) | 42 (42%) | 0.66 |
| **% PTP (IQR)** |  |  |  |  |
| 2013 | 44.0 (24–58.0) | 37.0 (20–56.0) | 44.0 (28–65) | 0.014 |
| 2019 | 14.0 (10–26.0) | 13.0 (6–24.0) | 22.0 (11–32) | <0.001 |
| **PTP AD-F 2013** |  |  |  |  |
| Yes < 15% | 34 (8.3%) | 33 (10.7%) | 1 (1%) | <0.001 |
| 15–85% | 350 (85.8%) | 261 (84.7%) | 89 (89%) |  |
| >85% | 24 (5.9%) | 14 (4.5%) | 10 (10%) |  |
| PTP 2019 |  |  |  |  |
| 1–5% | 43 (10.6%) | 43 (14.0%) | 0 (0%) | <0.001 |
| 5–15% | 165 (40.6%) | 129 (42%) | 36 (36%) |  |
| 15–85% | 195 (48.0%) | 133 (43.3%) | 62 (62%) |  |
| >85% | 3 (0.7%) | 2 (0.7%) | 1 (1.0%) |  |

Abbreviations: CAD = coronary artery disease; PTP = pre-test probability; AD-F = Diamond Forrester

Figure 3 illustrates the observed and expected prevalence of disease across probability quartiles. In the case of the PTP2013 model, there was a marked discrepancy in frequencies, with predicted probabilities exceeding observed values, particularly in the upper two quartiles. Conversely, for the PTP2019 model, observed probabilities were higher than predicted values, especially in the first two quartiles.

### Discrimination

The discrimination ability of both models was similar, with the PTP 2019 model showing a slightly lower C-statistic of 0.610 (95% Confidence Interval [CI]: 0.544–0.676) compared to 0.633 (95% CI: 0.570–0.696) for the PTP 2013 model. Analysis of the ROC curves revealed no statistically significant difference between the two models (p = 0.060). The effect size, measured by Cohen’s delta for the area under the curve (AUC), was 0.396 for PTP 2019 and 0.481 for PTP 2013 (see Figure 4).

**Figure 3.**
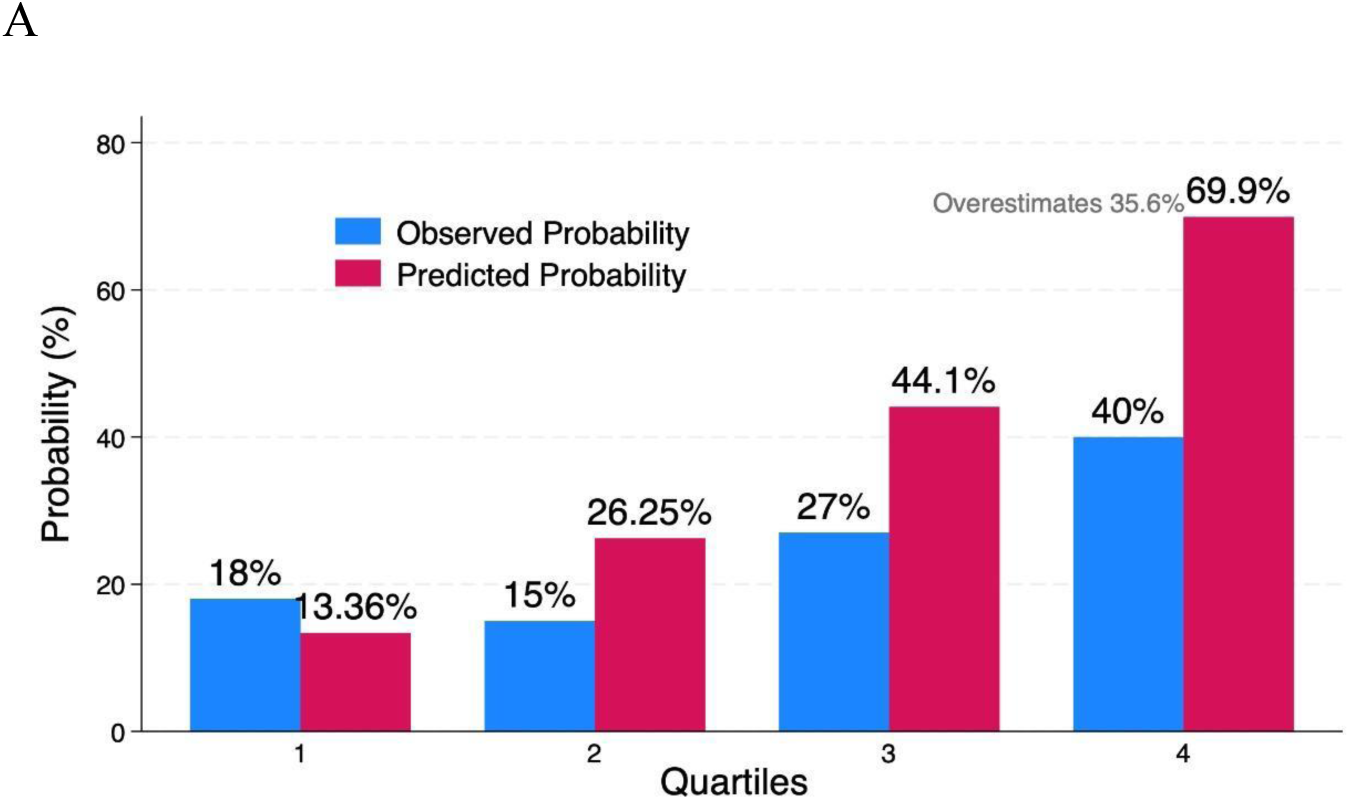
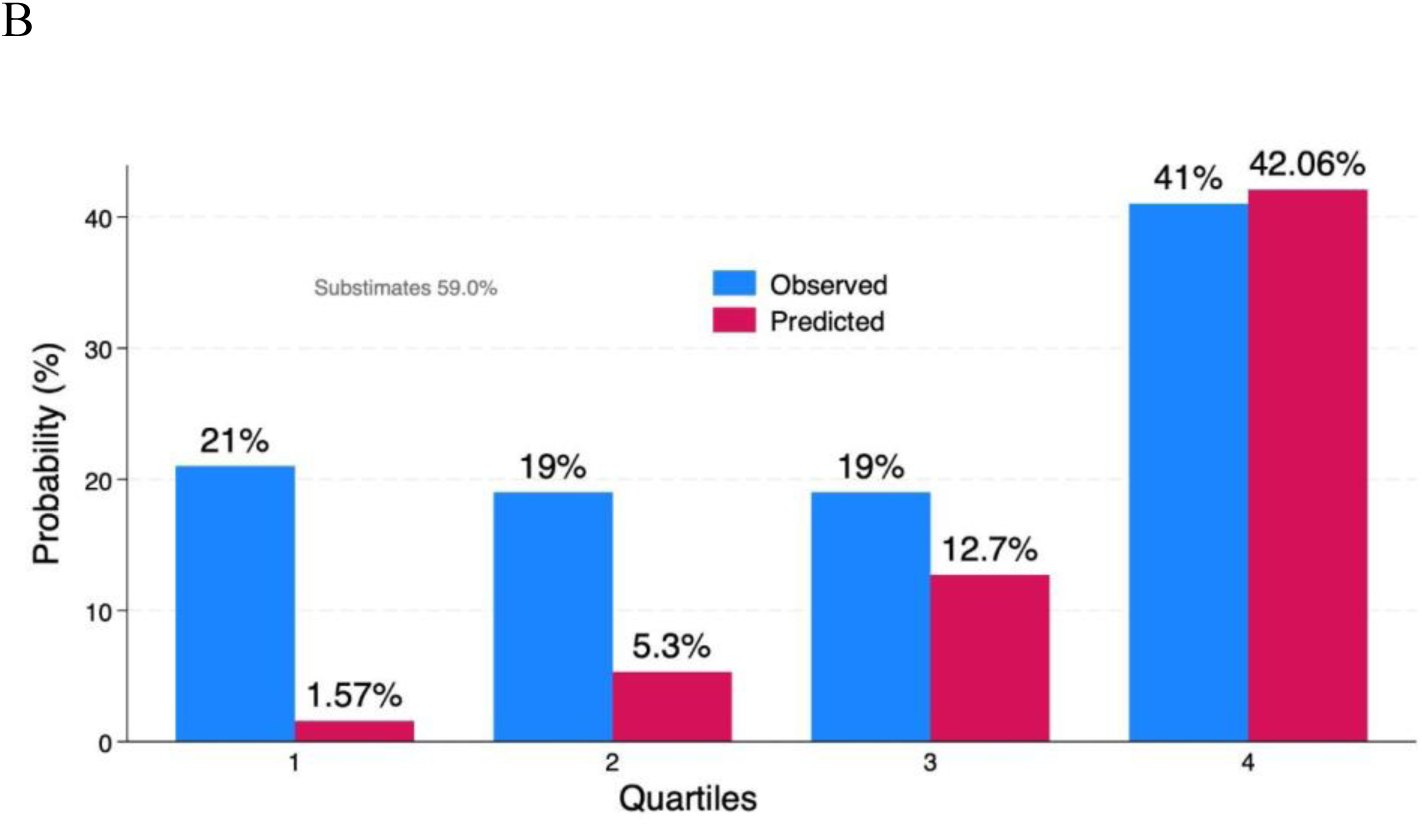
Observed and expected prevalence of disease across probability quartiles. (A) PTP 2013; (B) PTP2019

**Figure 4.**
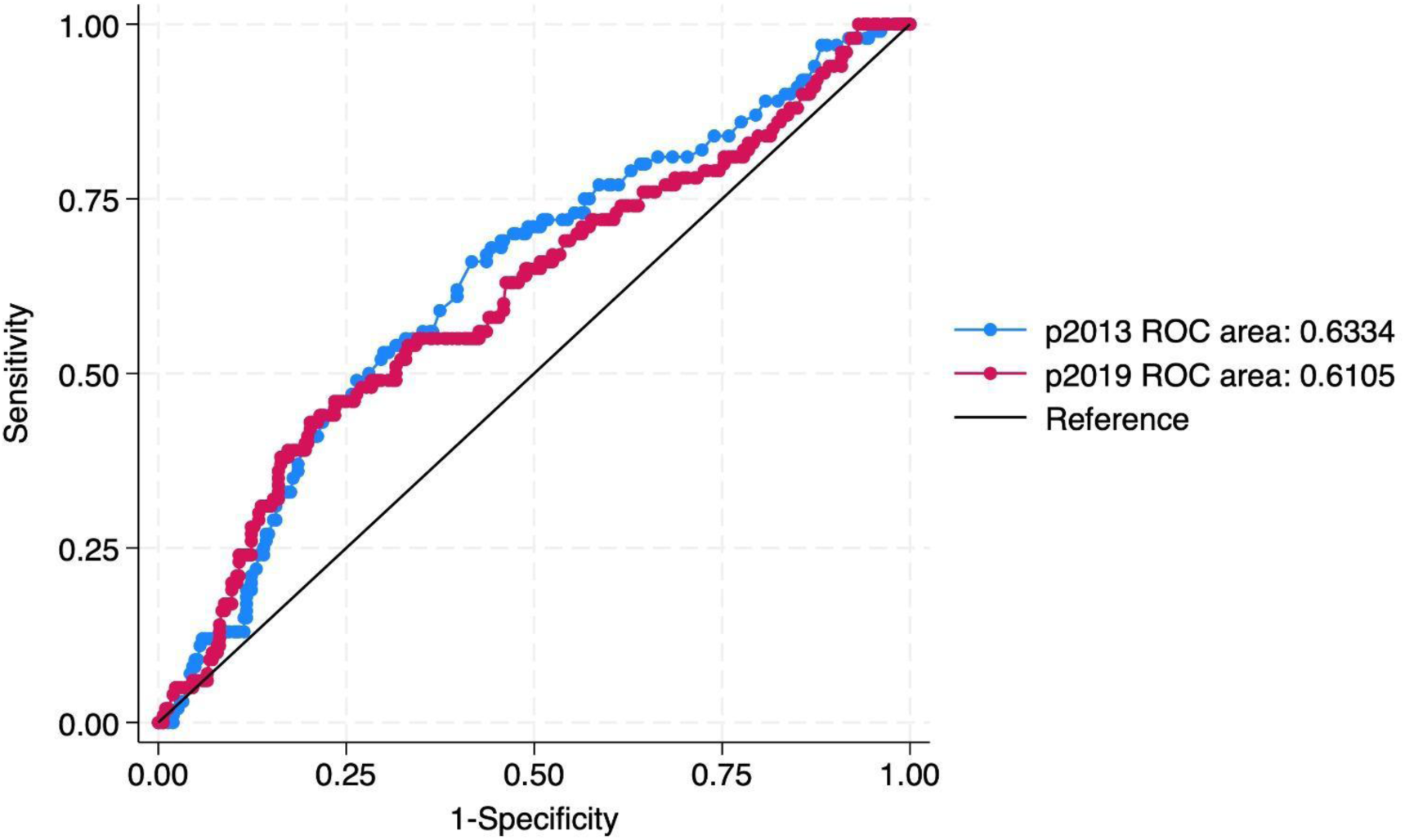
Receiver Operating Characteristic (ROC) Curve Comparing the Area Under the Curve for the 2013 (blue) and 2019 (red) Pre-Test Probability Models from the European Society of Cardiology.

### Net Reclassification Improvement

Reclassification performance was assessed using the Net Reclassification Improvement (NRI) metric. Both probability models were categorized into three risk groups based on cut-offs: low probability (<5%), intermediate probability (5–15%), and high probability (>85%) (Table 3). Using the PTP 2013 model as the reference, the NRI was 0.1474 (95% CI: 0.0342–0.258), with a p-value of 0.062, indicating a non-significant 14.7% of individuals were correctly reclassified by the PTP 2019 model. Table 3 details the number of individuals classified within each probability category by both models, as well as the overlap between classifications. Overall, 47% of individuals were assigned to the same risk category by both models. Notably, among the 333 individuals classified as intermediate probability by the PTP 2013, 52.7% were reclassified to the low probability group according to the PTP 2019.

**Table 3.**
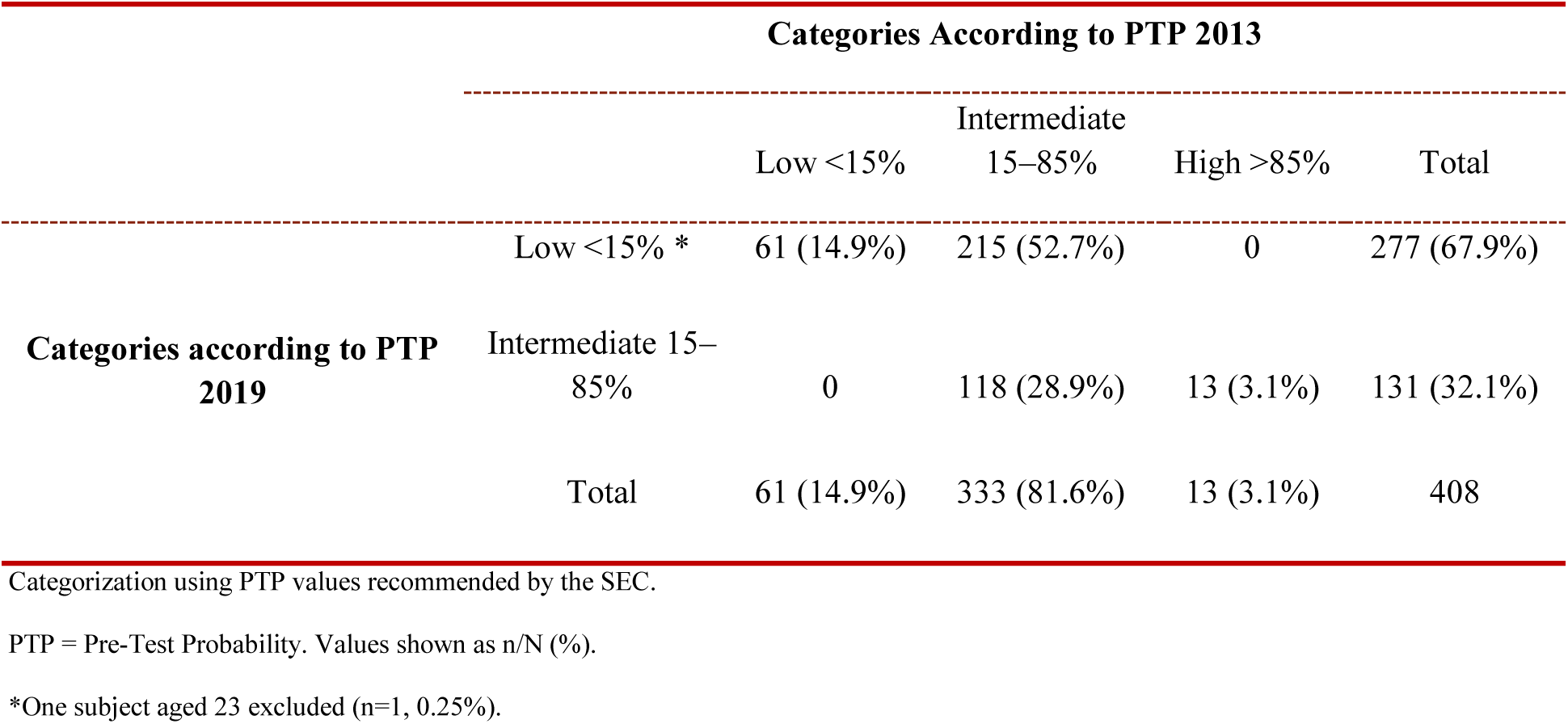
Reclassification of Patients.

|  |  | Categories According to PTP 2013 |  |  |  |
| --- | --- | --- | --- | --- | --- |
|  |  | Low <15% | Intermediate<br>15–85% | High >85% | Total |
| Categories according to PTP<br>2019 | Low <15% * | 61 (14.9%) | 215 (52.7%) | 0 | 277 (67.9%) |
|  | Intermediate 15–<br>85% | 0 | 118 (28.9%) | 13 (3.1%) | 131 (32.1%) |
|  | Total | 61 (14.9%) | 333 (81.6%) | 13 (3.1%) | 408 |

Categorization using PTP values recommended by the SEC.
PTP = Pre-Test Probability. Values shown as n/N (%).
\*One subject aged 23 excluded (n=1, 0.25%).

### Clinical Utility

In the validation cohort, both PTP models were calculated, and a comprehensive sensitivity analysis was performed using a 15% probability cut-off (Table 4). The PTP 2019 model identified 67% of patients (n = 276) as low probability; among these, 52 patients were found to have disease, yielding a sensitivity of 48% (95% CI: 37.9–58.2) and a positive predictive value (PPV) of 36% (95% CI: 28.4–45.5). In contrast, the PTP 2013 model reported a higher sensitivity of 90% (95% CI: 82.4–95.1). Both models achieved a negative predictive value (NPV) exceeding 80%. The PTP 2019 model did not predict any probabilities above 85%, and both models demonstrated low discriminatory power according to the diagnostic odds ratio (DOR).

## DISCUSSION

### Main Findings

This study performed the external validation and comparison of two clinical prediction models for coronary artery disease (CAD) in a Colombian cohort. In line with previous research, it was reported that the PTP 2013 model overestimates the probability of coronary artery disease. (Genders et al., 2018; Reeh et al., 2019). On the other hand, in our study this finding is possibly related to a higher prevalence, being 25% for CAD, which was higher than that reported in contemporary cohorts. (Juarez-Orozco et al., 2019b & Winther et al., 2022b) 14.9 and 8% respectively.

The proportion of CAD cases in the present study was 25%, a value that is in line with those reported in clinical guidelines previously published by Montalescot et al. (2013), where a prevalence of 44.5% was documented. However, recent cohorts have been showing a progressive decrease in prevalence, reaching figures as low as 8% according to the findings of Winther et al. (2020, 2022). This substantial decline in prevalence could be explained by both epidemiological and methodological factors, including lifestyle, preventive therapies, new patterns of clinical presentation, and a greater inclusion of patients undergoing coronary computed tomography angiography. It is noteworthy that the cohorts used to generate and validate the 2013 PTP models (Genders et al., 2011) and PTP 2019 (Juarez-Orozco et al., 2019a) were developed exclusively in patients from European and American populations. However, a recent Asian cohort (Baskaran et al., 2024), performed an external validation of the model, reported a prevalence very similar to that identified in our study. This situation highlights the great importance of validating these prediction models in different populations and even the need to adjust probabilities according to local clinical and epidemiological characteristics.

**Table 4.** Clinical utility of 2 categorized PTP models based on probability.

| Predicted Probability | PTP 2013 |  |  | PTP 2019 |  |  |
| --- | --- | --- | --- | --- | --- | --- |
|  | Total population<br>n=408 | With CAD | Without CAD | Total population<br>n=408 | With CAD | Without CAD |
| Low <15% | 61 (14.7%) | 10 (2.4%) | 51 (12.5%) | 276 (67.8%) | 52 (12.7%) | 225 (55.1%) |
| Intermediate 15-85% | 333(81.8%) | 97 (23.7%) | 297 (72.7%) | 131 (32.1%) | 48 (11.7%) | 83 (20.3%) |
| High >85% | 13 (3.1%) | 3 (0.7%) | 11 (2.7%) | 0 (0%) | 0 (0%) | 0 (0%) |
| <15% vs. >15% |  |  |  |  |  |  |
| Sensitivity |  | 90% (82.38 -95.10%) |  |  | 48% (37.9-58.22%) |  |
| Specificity |  | 16.56% (12.58 - 21.19%) |  |  | 73% (67.73 - 77.93%) |  |
| PPV |  | 25.94% (21.40-30.89%) |  |  | 36.64% (28.40-45.50%) |  |
| NPV |  | 83.61% (71.91-91.85%) |  |  | 81.23% (76.12-85.65%) |  |
| LR+ |  | 01.08 |  |  | 1.78 |  |
| LR- |  | 0.60 |  |  | 0.71 |  |
| DOR |  | 1.79 |  |  | 2.5 |  |

PPV = positive predictive value; NPV= negative predictive value; LR+ = positive likelihood ratio; LR- = Negative likelihood ratio; DOR = diagnostic odds ratio

Similar to recent studies, the prevalence of typical chest pain found in our study is similar between men and women, although with some different characteristics (Hemal et al., 2016). While the pooled analysis of three European cohorts identified atypical angina as the predominant symptom (Juarez-Orozco et al., 2019; Min et al., 2007), in our investigation we found that non-anginal pain was more frequent (47%), regardless of gender (Sekhri et al., 2016; Genders et al., 2011), constituting 44% of patients with CAD. This discrepancy could be related to differences in risk factors, as patients with non-anginal chest pain in our population also had higher rates of diabetes, hypertension, smoking, kidney disease, obesity, and a family history of coronary artery disease. Despite these variations in presentation, chest pain remains the cardinal symptom of coronary artery disease.

Dyspnea has acquired greater clinical relevance as it has been recognized as an isolated initial symptom associated with exercise or physical exertion (Douglas et al., 2023), as well as a prognostic factor in stable angina (Abidov et al., 2005). Our findings corroborate this clinical importance, as we observed a high prevalence of dyspnea as a clinical manifestation.

In our study, when comparing the PTP 2013 and PTP 2019 models, we observed similar discrimination patterns, which could be explained by the use of the same variables determined by the European Society of Cardiology (Vrints et al., 2024). Alternative models with greater discriminatory power exist, favored by the measurement of the Agatson coronary calcium score (CACS) on CT (Winther et al., 2019, 2020). The CACS is recognized as the most relevant predictor in terms of predicting coronary obstruction compared with clinical characteristics and CAD risk factors used in other models (Genders et al., 2012), however, the use of this factor implies the performance of a high-cost noninvasive diagnostic test. In particular, Colombia faces structural barriers in health (Bernal et al., 2012), such as limited access to advanced medical technology, especially in rural areas and primary care centers, which makes it difficult to systematically apply it as a strategy to predict the probability of CAD at the first level of care.

Recent studies document a clear trend toward downward reclassification of coronary risk, as reported by Foldyna et al. (2019), where a 50% reclassification of patients from intermediate to low probability was observed. Another study shows a reclassification from low to very low probability (Winther et al., 2020b). These findings should be interpreted with caution considering that these cohorts had a low prevalence of coronary artery disease, possibly due to selection bias, since they included only patients referred for CT angiography. Our study, by including both patients undergoing CT angiography and coronary angiography (CA), allowed us to assess the full spectrum of CAD. It also reported a reclassification of 64% of patients from intermediate to low probability. Despite this, the net improvement in reclassification was not significant.

### Clinical Implications of 2019 PTP Model

Determining the optimal threshold in probability models for CAD represents a contemporary epidemiological challenge of considerable clinical relevance. Therefore, establishing an accurate cutoff point involves crucial decisions such as the prescription of specialized diagnostic tests and the subsequent patient management. Following current recommendations (Vrints et al., 2024), we adopted a 15% cutoff point to define low probability in our study. By comparatively evaluating the 2013 PTP and 2019 PTP models, we identified relevant findings for their application in our population.

When analyzing the diagnostic performance of both models, our findings are consistent with the reported external validation of a Portuguese cohort (Lopes et al., 2022), where both models exhibited comparable discriminatory power. However, the clinical application of the model will initially be carried out in primary care services. In this sense, the predictive model prioritizes the correct identification of patients with the disease, making it preferable to use a more sensitive than specific model (Trevethan, 2017; Usher-Smith et al., 2016). Therefore, the 2013 ESC-PTP model, which reports a sensitivity of 90% (CI 82.38– 95.10%), is appropriate compared to the 2019 ESC-PTP model, which reports a sensitivity of 48% (IC 37.9-58.22%).

The 2019 PTP model, although slightly more accurate overall (as reflected by the DOR), introduces a clinically unacceptable risk by generating approximately 13% false negatives. Among these, patients classified as having ’low probability’ would be managed expectantly, without the need for functional cardiac testing—leading to underdiagnosis of CAD. These findings are consistent with those observed in another external validation study conducted in a Chinese population (Zheng et al., 2024).

The underdiagnosis associated with the 2019 PTP model could compromise the timely inclusion of patients in disease-modifying programs, such as lifestyle changes and comprehensive evaluation of cardiovascular risk factors (Trevethan, 2017). Additionally, it may negatively impact populations with high-risk factors or comorbidities such as familial hypercholesterolemia, obesity, among others. These populations are associated with a higher prevalence of CAD—factors that are not adequately accounted for in this model (Reeh et al., 2019b).

Recent studies report a low annual risk (1.1%) of major cardiovascular events in the low-probability category (Bjerking et al., 2022), supporting the use of expectant management. However, these results come from populations with markedly different characteristics and prevalences of CAD. Notably, the absence of similar studies in Latin America raises concerns about the medium- and long-term cardiovascular safety of this approach in our population.

### Strengths and Limitations

There is currently no validated PTP model for CAD specifically designed for the Latin American population. Our study represents an important first step by evaluating the external validity of the PTP 2013 and PTP 2019 models in a Colombian cohort. A methodological strength was the inclusion of patients who underwent non-invasive cardiac testing and were subsequently referred for coronary angiography. This allowed the inclusion of patients initially classified as intermediate probability, covering the full clinical spectrum of CAD—a feature that enhances the validity of our findings and their potential applicability in the regional context.

Although most participants were referred from coronary angiography, a strategy was implemented to mitigate verification bias by including patients who underwent coronary CT angiography, thereby incorporating individuals with low pre-test probability. While the single-center design is a methodological limitation, it is noteworthy that the data were extracted from reliable institutional records. This database includes patients with diverse demographic characteristics, as the institution serves as a referral center for multiple regions of the country, which increases the variability of the sample and its population representativeness.

Looking ahead, it is imperative to develop a contemporary cohort that prospectively validates clinical probability for coronary artery disease. We propose a comparison between existing models and the newly recommended risk factor-weighted clinical likelihood (RF-CL) model, as outlined in the latest European Society of Cardiology guidelines (Vrints et al., 2024)

## CONCLUSION

In our Colombian cohort, the 25% prevalence of CAD was notably higher than that reported in large contemporary European and North American cohorts, a finding likely attributable to differences in the epidemiological profile and clinical characteristics of our population. The methodological challenge of identifying the predictive model with the best diagnostic performance for our setting remains. Although both models evaluated (PTP-2013 and PTP-2019) demonstrated limited discriminatory capacity, there were differences in their clinical applicability

In conclusion, considering the sociodemographic and clinical characteristics of the studied cohort, the PTP-2013 model is preferred due to its higher sensitivity—despite overestimating CAD—compared to the PTP-2019 model, which underestimates CAD and leads to a concerning level of underdiagnosis. Ideally, a predictive model should be developed within our population context and subsequently compared with the most recently validated prediction models.

## Data Availability

Data will be available to access.

https://doi.org/10.5281/zenodo.17518364

## Annex 1

Comparison of the proportion of observed data with predicted probabilities in accordance with the ESC-PTP 2013 model and the ESC-PTP 2019 model depending on sex. Women n=197 and Men n=211. Women ESC-PTP 2013 (A); Women ESC-PTP 2019 (B); Men ESC-PTP 2013 (C); Men ESC-PTP 2019(D).

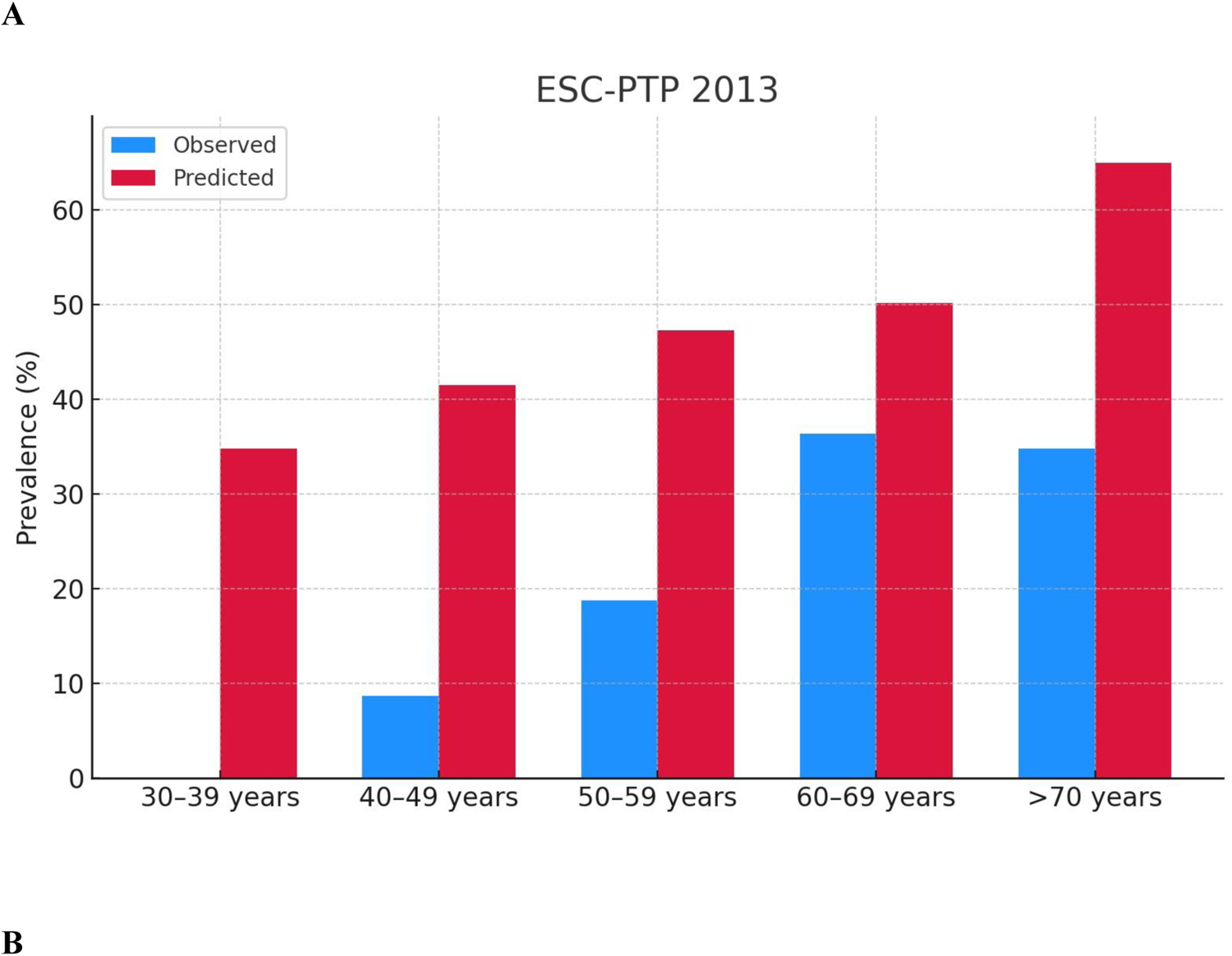
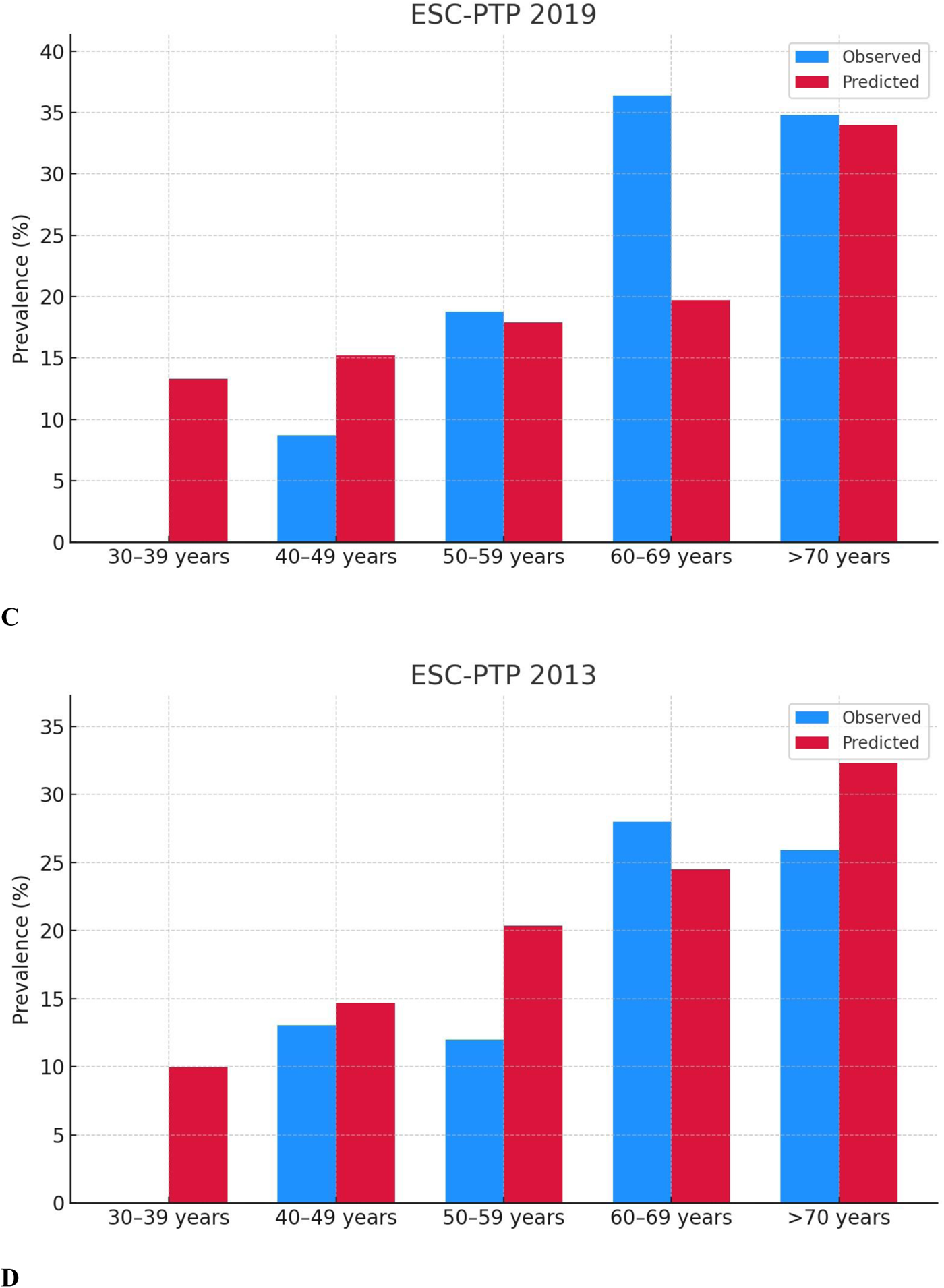
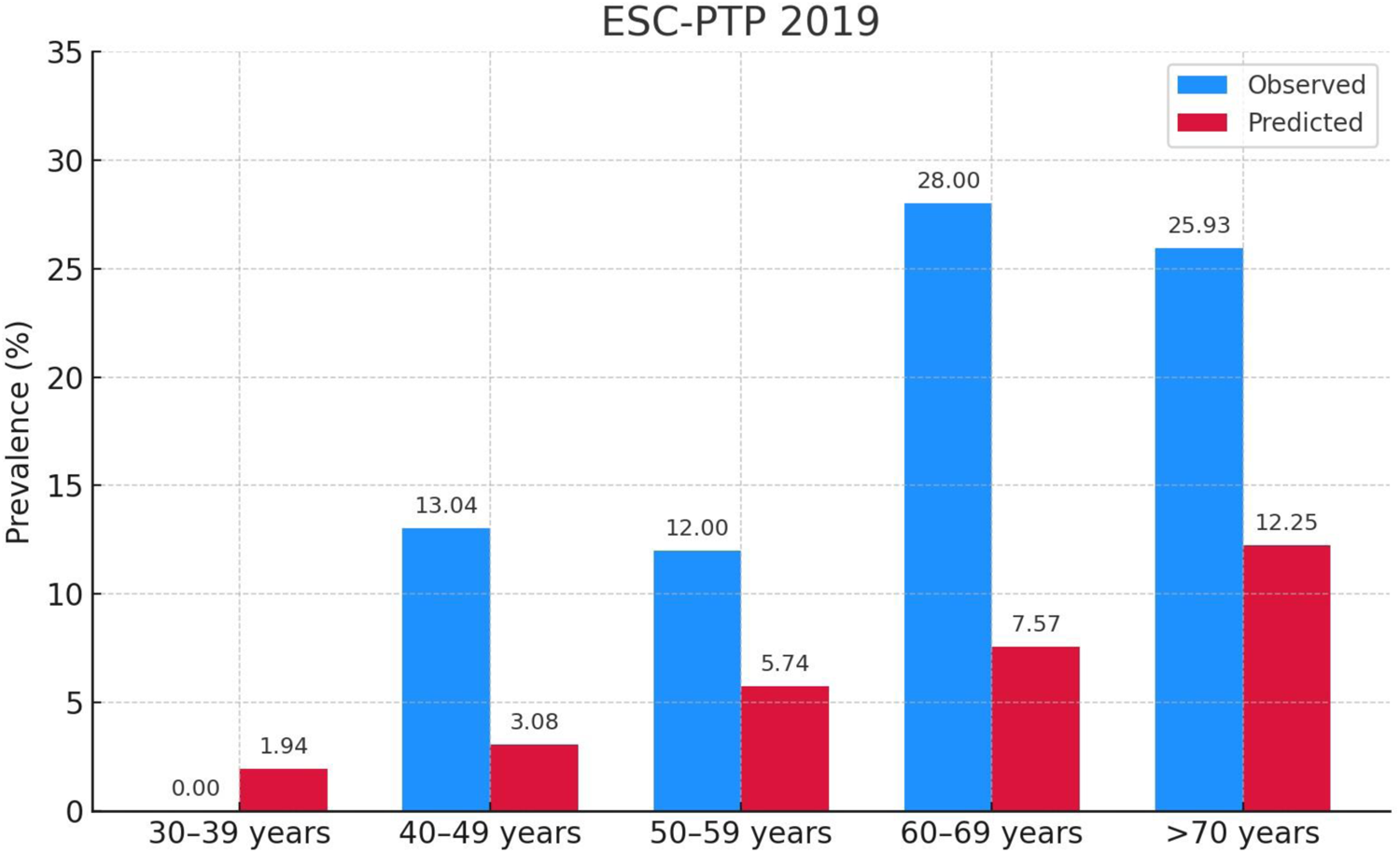

